# Socio-demographic, clinical, hospital admission and oxygen requirement characteristics of COVID-19 patients of Bangladesh

**DOI:** 10.1101/2020.08.14.20175018

**Authors:** Ratindra Nath Mondal, Md. Abdur Razzak Sarker, Anupam Das, S. M. Ahsanul Kabir Ahsan, Shah Md. Sarwer Jahan, Aklima Sultana, Jafrin Sultana, Moni Rani

**Author notes:** **Correspondence** Dr. Ratindra Nath Mondal President, Society of General Physician Founder-Daktarkhana (GP center), Rangpur, Bangladesh, Email;.

## Abstract

**Background:** Clinical presentation, hospital admission and outcome of COVID-19 pandemic are different from one country to another. So every country should have their own data regarding COVID-19.

**Subjects and Methods:** This was an online cross-sectional survey, carried out in RT-PCR positive COVID-19 adult patient. A preformed questionnaire adapted in Google form and circulated through online to collect data. Informed consent was ensured before participation in this study.

**Results:** We have studied of 305 RT-PCR positive COVID-19 patients, mean age was 36.32 (+/− 12.369) years with male predominance and majority were doctor 46.9%. 48.8% of the patient’s family members affected along with them. After COVID-19 pandemic 31.7% did not go out of their house. For prevention 72.4%)of the patients used mask and 38.8% used hand gloves during outing. 28.7% reported to be affected while working in hospital, 21.8%were affected in their office. Fever (80.1%), cough (57.7%), pain in throat (50.8%), rhinitis (45.9%) and loss of smell (45%) & taste sensation (45.6%) were the most common symptoms. Sample was given average 3 (+/− 3.384) days after onset of the symptoms and report was delivered average 4.57 days after giving sample. After getting result 32.1% of the patients seek treatment in telemedicine and 23% directly consulted in Government hospital, only 2% reported to be treated by the Government designated telephone number. 73.1% of the patient took both steam inhalation and warm water gargling. Paracetamol and antihistamine was the most commonly use drugs 69.8% and 71.5% respectively, besides 47.5% took ivermectin, 41.6% took azithromycin, 35.1% steroid, 34.8% took doxycyclin and 21.6% hydroxychloroquine. Among the patients only 20.3% needed hospital admission. The patients admitted in hospital average 5 (+/-3.922) days after onset of symptoms and hospital staying was 9.2 days (average). Breathless (54.83%) was the major cause of hospital admission and 19.35% patients admitted due to fear. 14% patients needed oxygen and average duration of oxygen was 4.84 days. 81.3% patients has taken oxygen in hospital and 18.8% at their home. Among the patient 2.2% needed ICU and artificial ventilation needed for 1.1% patients. The patients recovered after 17.74 days (average) from the onset of symptom. COVID-19 became negative (RT-PCR) at15^th^ day from onset of symptom. 91.8%) were in mental stress to become the cause of infection spread to other family members, 20.7%) reported that they were anxious that they would not get oxygen or ICU if required and 27.9% were suffering from fear of death. Weakness was the most common post disease symptom in 57.4% cases. Average time required for coming back to normal life was 21.59 days (+/-7.901) ranging a wide range from 5 to 60 days.

**Conclusion:** COVID-19 patients were mostly male, health worker. Fever, cough, pain in throat were most common symptoms. Hospital admission required in only one-fifth cases and ICU required for only 2% patients. Weakness was the most common post disease symptom.

## Introduction

In early December 2019, the first pneumonia cases of unknown origin were identified in Wuhan, the capital city of Hubei province.^1^ The pathogen has been identified as a novel enveloped RNA betacoronavirus.^2^ The virus was initially designated as the novel coronavirus (2019-nCoV), but after the global agreement, it was renamed Corona Virus Disease 2019 (COVID-19).^3^ The World Health Organization (WHO) has declared coronavirus disease 2019 (Covid-19) a public health emergency of international concern (pandemic) in 11 March, 2020.^4^ At the time of writing this article (3^rd^ august) globally total 18,258,528 cases has been identified and 693,395 patients died.^5^ In Bangladesh first case has been declared^6^ in 8^th^ March, 2020 and by early August total 242,102 cases and 3,184 deaths occurred in this country.^5^ The clinical spectrum of COVID-19 varies from asymptomatic or paucisymptomatic forms to clinical conditions characterized by respiratory failure that necessitates mechanical ventilation and support in an ICU to multiorgan and systemic manifestations in terms of sepsis, septic shock, and multiple organ dysfunction syndromes (MODS). In one of the first reports on the disease, Huang et al. illustrated that patients (n. 41) suffered from fever, malaise, dry cough, and dyspnea. Chest computerized tomography (CT) scans showed pneumonia with abnormal findings in all cases. About a third of those (13, 32%) required ICU care, and there were 6 (15%) fatal cases.^1^ The disparity of^1, 3, 7, 8, 9, 10^ sociodemographic characteristics between countries is also obvious and therefore, the management strategy of one country needs to be individualized. During the initial phase of the Covid-19 outbreak, the diagnosis of the disease was complicated by the diversity in symptoms and imaging findings and in the severity of disease at the time of presentation.

Therefore this study was carried out to determine the socio-demographic and clinical characteristics of the COVID-19 patients of our country.

## Methodology

This was an online cross-sectional survey, carried out by the Research wing of Daktarkhana (GP center of Bangladesh), among the RT-PCR positive COVID-19 adult patients (aged >18 years) who are using Facebook (the most widely used social media of the country) and Twitter, Viber, WhatsApp etc. A recruitment post was posted in the Facebook news feed of the authors. This was subsequently shared by the whole author team in Twitter and various groups in “Facebook Messenger”, “WhatsApp”, and “Viber” through their acquaintances. This post contained a brief introduction about the background, objective, procedures, voluntary nature of participation, declarations of anonymity and confidentiality, and notes for filling in the questionnaire as well as the link to the online questionnaire (Google form).

Persons of Bangladeshi nationality understood the content of the post and agreed to participate in the study were instructed to complete the questionnaire by clicking the link. In addition, the questionnaire link or link of Google form was distributed by the friends and family of the authors through Short Message Services (SMS) to reach as many respondents as possible. The Google form consists of five parts: a brief introduction, consent statement, demographic profile, clinical and radiological information, and Patient Health Questionnaire (PHQ-9).

A contact email address and phone number were provided in the post to contact regarding any difficulty in completing the form. Initially, pretesting of the questionnaire was performed among 10 random participants, and the experience of the piloting was used to make a final adjustment before the final Facebook post was published online. The form was set so that if any person clicked the link to participate and did not agree with the informed consent statements, he or she could not proceed to the next sections of the questionnaire.

### Ethics statement

Before the commencement of the study, formal ethical approval was obtained from the Ethical Review Committee (ERC) of Hypertension and Research Center, Rangpur, Bangladesh. All methods were performed in accordance with the current Declaration of Helsinki. All participants gave informed written consent before participation.

### Data cleaning and analyses

Due to automation of the Google form, fill up data were recorded into the Google drive as sheets in ‘comma separate value’ (csv) format. The sheet was cleaned, organized and imported into the statistical software SPSS 20 (SPSS Inc, Chicago, IL, USA) for final analysis. Descriptive statistics were used during analysis, where continuous variables were expressed as the mean±standard deviation and categorical variables were expressed as count (percentage).

## Result

In this study, we have studied a total of 305 COVID-19 patients of both sexes. Male (205) were more than female (100) ratio is (2:1). Mean age of the patient was 36.32(+/-12.369) years. (Table 1 shows the socio-demographic characteristics of the study population).

**Table 1.**
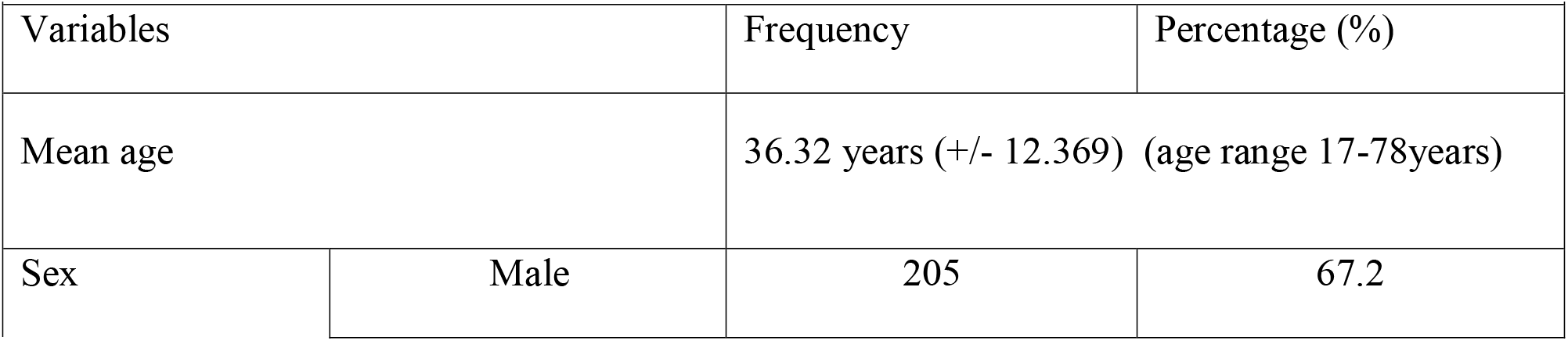
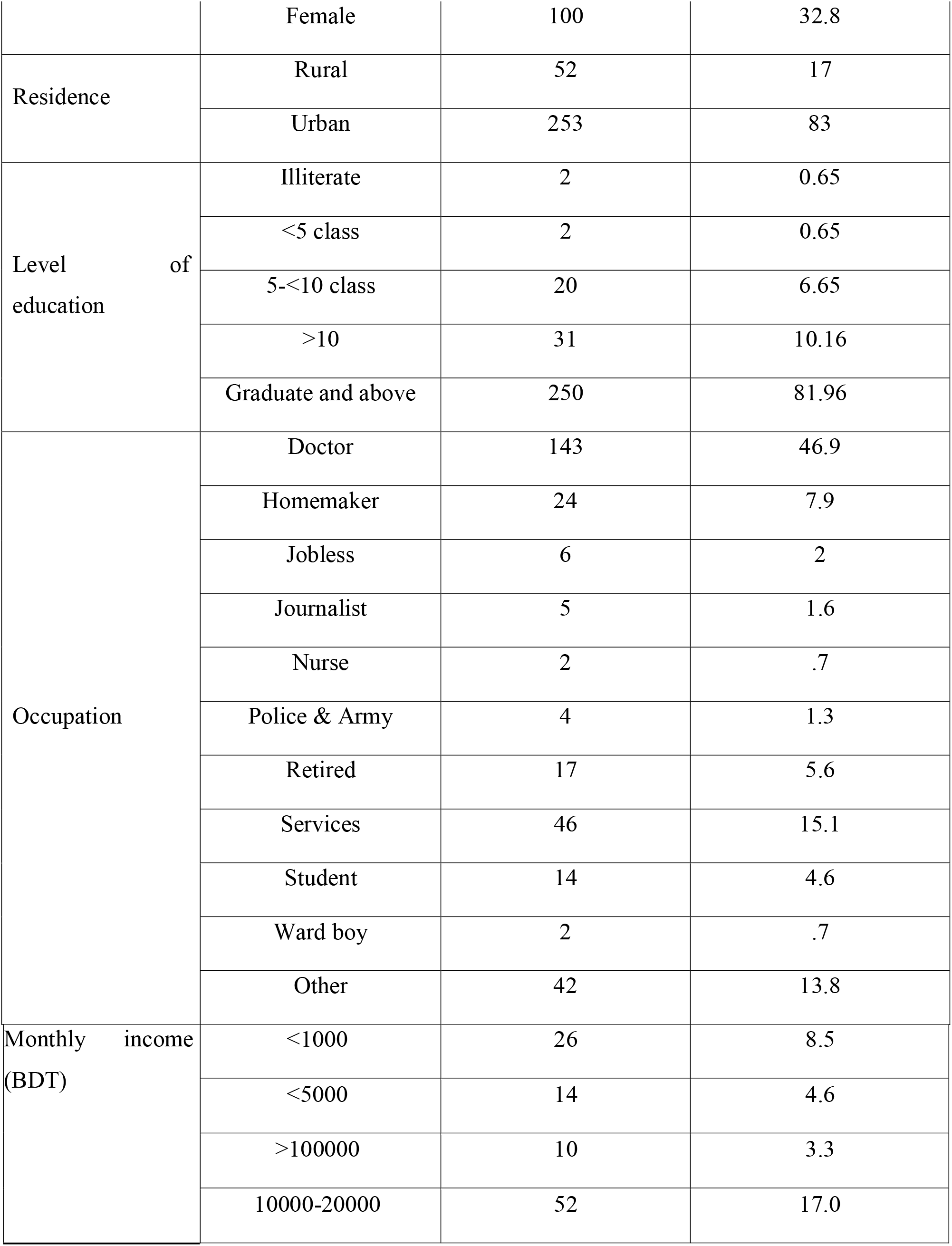
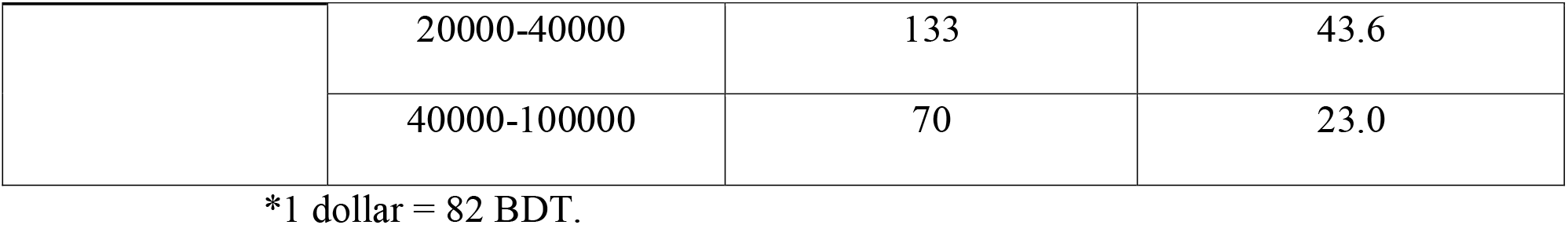
shows the socio-demographic characteristics of the study population

Maximum (n=307, 96.8%) patients had no history travelling to foreign country immediately before COVID-19 outbreak in Bangladesh. After covid-19 outbreak in Bangladesh 159 (38.8%) patients used to go to office regularly, 31 (7.6%) use to go for buying of daily needs and 130 (31.7%) did not go out of their house. For prevention of COVID-19, 297 (72.4%) of the patients used mask and 159 (38.8%) used hand gloves during outing. When returning to home washed their hands (n=135, 32.9%) and cloths (n=119, 29.9%) with soap. Table II shows the protective measure taken by the patients when went to outside and returning home.

**Table II:** Measures to prevent COVID-19 in outdoor and after returning home

|  |  |  |  |
| --- | --- | --- | --- |
| Measures during outing |  | Measures after returning to home |  |
| Measures | Frequency/Percentage | Measures | Frequency/Percentage |
| Mask | 297 (72.4%) | Washing of all clothes | 119 (29.9%) |
| Social distancing | 247 (60.2%) | Hand washing with soaps | 135 (32.9%) |
| Hand gloves | 159 (38.8%) | Combination of the above | 188 (45.9%) |
| Nothing | 2 (0.5%) | None of the above | 4 (1%) |

On an average every patient has 4.75(+/− 2.1) family members, 48.8% family members affected concomitantly. Along with the patient average 2.26 (+/-1.287) family members affected concomitantly. Hypertension (n=59, 19.2%) was the commonest co-morbidity among the patients.

**Table III.** shows the co-morbidity of the patient.

| Table III showing the co-morbidity of the patient |  |
| --- | --- |
| Co-morbidity | Frequency/Percentage |
| Hypertension | 59 (19.2%) |
| Asthma | 54 (17.6%) |
| Diabetes mellitus | 42 (13.7%) |
| IHD | 15 (4.9%) |
| Stroke | None |
| Kidney disease | 6 (2%) |
| Liver disease | 5 (1.6%) |
| Others | 25 (8.1%) |
| No chronic disease | 99 (69.4%) |

According to the suspicion of the patient major bulk 28.7% reported to be affected while working in hospital, 21.8%were affected in their office and 12.4% affected from direct contact with COVID-19 positive family members in their home. Table IV shows details.

**Table IV.** showing where the patients thought to be affected

| Table IV showing where the patients thought to be affected |  |
| --- | --- |
| Where the patients thought to be affected | Frequency/Percentage |
| At hospital while giving treatment to patients | 88 (28.7%) |
| At office | 67 (21.8%) |
| At home from affected family members | 38 (12.4%) |
| Public transport and Public Places | 8 (3%) |
| At hospital while treatment purpose of family members | 9 (2.9%) |
| No idea | 95 (31.14%) |

Fever (80.1%), cough (57.7%), pain in throat (50.8%), rhinitis (45.9%) and loss of smell (45%) & taste sensation (45.6%) were the most common symptoms. Table V showing the symptoms of the patients. There was no symptom only in few cases 8 (2.62%).

**Table V.** showing the symptoms of the patients

| Table V showing the symptoms of the patients |  |
| --- | --- |
| Symptoms | Percentage (frequency) |
| Fever | 246 (80.1%) |
| Weakness | 224 (73%) |
| Cough | 177 (57.7%) |
| Pain in throat | 156 (50.8%) |
| Body ache | 155 (50.5%) |
| Loss of appetite | 153 (49.8%) |
| Rhinitis | 141 (45.9%) |
| Loss of taste | 140 (45.6%) |
| Loss of smell | 138 (45%) |
| Diarrhea | 83 (27%) |
| Breathlessness | 72 (23.5%) |
| Chest pain | 52 (16.9%) |
| Frequent micturition | 31 (10.1%) |
| Vomiting | 31 (10.1%) |
| Redness of the eyes | 28 (9.1%) |
| Watering from eyes | 10 (3.3%) |
| Skin rash | 10 (3.3%) |

230 (75.4%) of the patients has given sample for RT-PCR in hospital and remaining given at home. Sample was given average 3 (+/− 3.384) days after onset of the symptoms. Report was delivered average 4.57 days after giving sample (minimum in the day of report and maximum 27 days). After getting result 32.1% (n=98) of the patients seek treatment in telemedicine and 70 (23%) directly consulted in Government hospital, only 6 (2%) reported to be treated by the Government designated telephone number.

Among the non-pharmacological measures majority 73.1% (n=223) of the patient took both steam inhalation and warm water gargling. 10.8% (n= 33) used gargling and 4.6% (n=14) used stem inhalation; 9.50% (n=29) did not follow any of the non-pharmacological measures. 84.6% (n=258) of the patients took medicine according to the advice of MBBS doctor and 9.8% (n=30) from other medics, very few took drug according to the advice of their non-medic relatives (n=10, 3.3%) and based on social media and internet (1.6%, n=5) as well. Following medicine were taken by the patients during their illness (Table VI)

**Table VI:** Drugs taken by the patient during their illness

| Table VI: Drugs taken by the patient during their illness |  |  |
| --- | --- | --- |
| Name of the Drug | No of the patients took the Drug | Percentage of the patient took the drug |
| Paracetamol | 213 | 69.8% |
| Antihistamin | 218 | 71.5% |
| Ivermectin | 145 | 47.5% |
| Azithromycin | 127 | 41.6% |
| Doxycyclin | 106 | 34.8% |
| Hydroxychloroquine | 66 | 21.6% |
| Steroid | 107 | 35.1% |
| Rivarox | 64 | 21% |
| Inj. LMWH | 40 | 13.1% |
| Alternative | 7 | 2.3% |

Among the patients only 20.3% (n=62) needed to admit in hospital. The patients admitted in hospital average 5 (+/-3.922) days after onset of symptoms and on an average hospital stay duration was 9.2 days. 86% patients admitted in Government hospital and remaining admitted in private hospital. Breathless (54.83%) was the major cause of hospital admission, 19.35% patients admitted due to fear and 18% patients admitted according to the advice of their physician. 14% patients needed oxygen and average duration of oxygen was 4.84 days (minimum 01 day-15 days). 81.3% patients taken oxygen in hospital and 18.8% at their home. Among the patient 2.2% needed ICU and artificial ventilation needed for 1.1% patients. On an average the patients recovered after 17.74 days (minimum 2 days-60 days) from the onset of symptom. (Table VII shows details)

**Table VII:** Showing time to onset of improvement

| Table VII: Showing time to onset of improvement |  |
| --- | --- |
| Symptoms | Started to improve from onset of symptom |
| Fever | 7.38 +/- 6.39, Median 5, Mode 3, Range= 1-45 days |
| Loss of smell | 14.18+/- 8.057, Range= 3-45 days |
| Loss of taste | 3+/- 3.384, Mode 1, Range= 0-27 |
| Breathlessness | 9.5+/-9.237, Mode=7, Range: 1-45 days |

71.8% (n=219) patients has done their 2^nd^ RT-PCR test which is needed to declare negative. 2^nd^ test was done on average 15^th^ (14.93) days from onset of symptom and on 17.74^th^ (+/− 7.901) day they got the result that they became COVID-19 negative. After diagnosis majority 279 (91%) patients were isolated at their home and they broke the isolation 21.59 (+/-7.901) days after onset of symptoms. 280 (91.8%) were in mental stress to become the cause of spreading COVID-19 to other family members, 65 (21.3%) of the patient were in fear of never to become normal again, 63 (20.7%) reported that they were anxious they would not get oxygen or ICU if required, 85 (27.9%) were suffering from fear of death, 149(48.9%) were hopeful to become ok by time being and 111 (36.4%) reported they did not worry about anything. Weakness was the most common post disease symptom in 175 (57.4%) cases. Average time required for coming back to normal life was 21.59 days (+/-7.901) ranging a wide range from 5 to 60 days.

## Discussion

In our study we have studied of 305 RT-PCR positive COVID-19 patients. Mean age of the patient was 36.32(+/-12.369) years; another study^11^ from our country showed mean age was 42.59±14.43 years. Data from the Institute of Epidemiology, Disease Control, and Research (IEDCR)^10^ revealed that 42% of the Bangladeshi COVID-19 cases were aged between 21 and 50 years but in our study age range was 17-78 years. Age reflection is likely due to the most productive age of the life, for office/business they had to go out and thus they affected mostly. Higher age range is probably the affected older aged family members of the patient. Like other studies ^11, 7 12-14^ from home and abroad male were more than female in our study. MERS-CoV and SARS-CoV had a similar pattern of sex distribution.^13, 14^ Male predominance may be due to male are more mobile than female; outdoor activities like shopping, office mostly done by the male in our country. Besides it may be due to less case detection in female due to gender discrepancy. Ahmed et al (COVID-19 cases in Bangladesh) stated that the underreporting of female patients even after infection due to shyness or social stigma is a challenge to focus, which might be a potential source of rapid disease transmission.^15^

Ahmed et al. found 25.66% health workers among all COVID-19 cases of Bangladesh, ranking hospital physicians on top followed by nurses. Hasan et al.^1^ found 29.7% had healthcare-related jobs. In China^16^ 29% were healthcare workers. But in our study 48.3% patients were from hospital related job, majority of them were doctors (46.9%); this figure may be due to we have collected the data from public group and some doctor’s group. In spite of 72.4% patients used mask, 60.2% maintained social distancing and after returning to home 45.9% washed their hands and clothes, they were affected; this may be due to under quality of the mask or improper wearing of the mask. So for prevention of the COVID-19 quality and proper wearing of the mask needs to be ensured. Though 90.1% of the patients kept themselves isolated, 48.8% of their family members affected along with them and on average 2.26 (+/-1.287) family members were affected. So to protect family members isolation should be strictly maintained and the health care workers need to be quarantined in separate place before going to their family members.

Cardiovascular disorders (21%) and diabetes (16.3%) were the most prevalent co-morbidities among Covid-19 patients in a study of Iran.^17^ Previous studies have reported diverse prevalence rates ranging from 11% to 45% for cardiovascular disorders and from 13% to 35% for diabetes.^18-21^ A meta-analysis reports an overall pooled prevalence of 8.4% and 9.7%^22^ for cardiovascular disorders and diabetes, respectively, while another meta-analysis reports 12% and close to 8%.^23^ In our study hypertension (19.2%) was the most prevalent co-morbidity followed by Asthma (17.6%); DM was 13.7% and IHD was only 4.9%. Low prevalence of IHD may be due to younger age group of our study patients and IHD mainly developed in older age people; 91% patients were below 60 years age, 70.49% were below 40 years age. In our study most common symptoms were fever (80.1%), cough (73%), pain in throat (50.8%), bodyache (50.5% and loss of appetite (49.8%). In China symptoms of COVID-19 patient were fever (67.4%), dry cough (34.3%), and myalgia (28.5%).^24^ In a study of Iran fever (63.8%, cough (68.1%) and dysponea (72.7%) was the most common symptoms.^17^ In our study asymptomatic patient was 2.62%, this is due to most of our patients have done their test due to their symptoms.

A report from Bangladesh (Hasan et al^11^) found 87.8% of participants were symptomatic, wherein the most commonly reported symptoms were fever (77%), cough (56.8%), breathlessness 24.3%, myalgia (24.3%), sore throat (21.6%) and fatigue (17.6%). In our study breathlessness was present in 23.5%, which is much lower than WHO suggested^25^ prevalence (31-40%) and report from Iran.^17^ Loss of smell and taste sensation was present in 45% and 45.6% respectively, which may be distinguishing features of COVID-19. However among the non-respiratory symptoms diarrhoea was the most common (27%), followed by vomiting (10.1%), redness of the eyes (9.1%), watering from eye and skin rash was in 3.3%. Though telemedicine was not so popular for the patients before pandemic, in our study 32.1% patient consulted in telemedicine for treatment after confirmation of diagnosis. So, telemedicine may have an excellent prospect to provide treatment in future.

Although our national guideline for clinical management of COVID-19 promoted supportive and symptomatic treatment protocols along with judicial use of different modalities of drug regimen found to be effective by different trials.^26^ Majority 73.1% of the patient took both steam inhalation and warm water gargling; 47.5% took ivermectin, 41.6% took azithromycin, 35.1% steroid, 34.8% took doxycyclin and 21.6% hydroxychloroquine. The median time interval was 1.0 (IQR: 0-3.0) day from illness onset to the first medical visit, 0 (IQR: 0-2.0) day from the first medical visit to hospital admission, and 1.0 (IQR: 1.0-3.0) day from hospital admission to PCR confirmation.^24^ In our study RT-PCR was done on average 3 (+/− 3.384) days after onset of the symptoms. Report was delivered average 4.57 days after giving sample (minimum in the day of report and maximum 27 days). Only 20.3% needed to admit in hospital. The patients admitted in hospital average 5 (+/-3.922) days after onset of symptoms.

In earlier time of the COVID situation of Bangladesh Hasan et al.^11^ has shown that 6.8% patient needed oxygenation. But in our study 14% patients needed oxygen and 18.8% patients taken oxygen at their home. Oxygen therapy at home may be hazardous and patient may not get appropriate concentration of oxygen. So oxygen therapy must be discourages at home. Huang et al. found that 32% of patients needed admission to ICU, ^1^ where as 11.8% were in a study from Iran.^17^ In our study only 2.2% patients needed ICU admission and artificial ventilation needed for 1.1% patients. This small number may be due to the mutation of corona virus to less virulent type or better immunity of the Bangladeshi people. One study from China^27^ showed that 53.8% of the respondents had psychological impact as a consequence of outbreak and suffered with moderate or severe degree; 16.5% reported moderate to severe depressive symptoms; 28.8% reported moderate to severe anxiety symptoms, and 8.1% reported moderate to severe stress levels. Study from Bangladesh^11^ (Hasan et al.) found that 60% and 52.9% had anxiety and depression; among them 21.4% had severe anxiety and 8.1% had severe depression. In our study 91.8% were in mental stress to become the cause of spreading COVID-19 to other family members, 27.9% were suffering from fear of death, 20.7% were anxious that they will not get oxygen and ICU if they will require. After recovery more than half of the patients suffered from generalized weakness and average 21.59 days (+/-7.901) needed to come back to their normal life.

## Conclusion

COVID-19 patients were mostly male, health worker. Fever, cough, pain in throat were most common symptoms. Hospital admission required in only one-fifth cases and ICU required for only 2% patients. Weakness was the most common post disease symptom.

## Limitation

In our study data was collected from Facebook and some doctors group, so there was selection bias. Besides outcome of the patient was not addressed.

## Data Availability

Data available

https://docs.google.com/spreadsheets/d/1emc46KiJv6T_GDW83zyFCMRf6lC00Z-dPtUB2UnJqn8/edit#gid=2043165860

## Future direction

Hospital (both OPD and indoor) based or community based study with large sample size will be needed to obtain a full picture of the spectrum of clinical severity.

## Acknowledgement

The authors of this study are grateful to Most Happy Begum, moderators of GP practice guideline group and all the General physicians of Daktarkhana (GP centre).

